# An ultra-sensitive bacterial pathogen and antimicrobial resistance diagnosis workflow using Oxford Nanopore adaptive sampling sequencing method

**DOI:** 10.1101/2022.07.03.22277093

**Authors:** Hang Cheng, Yuhong Sun, Qing Yang, Minggui Deng, Zhijian Yu, Lei Liu, Liang Yang, Yu Xia

## Abstract

Metagenomic sequencing analysis has been implemented as an alternative approach for pathogen diagnosis in recent years, which is independent on cultivation and able to identify all potential antibiotic resistant genes. However, current metagenomic sequencing analysis methods have to deal with low amounts of prokaryotic DNA and high amounts of host DNA in clinical samples, which significantly decrease the overall microbial detection resolution. The recently released nanopore adaptive sampling (NAS) technology facilitates immediate mapping of individual nucleotides to a given reference as each molecule is sequenced. User-defined thresholds allow for the retention or rejection of specific molecules, informed by the real-time reference mapping results, as they are physically passing through a given sequencing nanopore. We developed a metagenomics workflow for ultra-sensitive diagnosis of bacterial pathogens and antibiotic resistance genes (ARGs) from clinic samples, which is based on the efficient selective ‘human host depletion’ NAS sequencing, real-time species identification and species-specific resistance gene prediction. Our method increased microbial sequence yield at least 8-fold in all 11 sequenced clinical Bronchoalveolar Lavage Fluid (BALF) samples (4.5h from sample to result), displayed 100% sensitivity and specificity for pathogen detection compared with cultivation method, and accurately detected antibiotic resistance genes at species level.

## Introduction

The traditional microbial diagnostic approaches for infectious diseases via conventional cultivation and antibiotic susceptibility test suffer from long reporting delays and low sensitivity and specificity[1, 2]. The implementation of metagenomic sequencing analysis (mNGS) has become a milestone for rapid diagnosis of infectious diseases, which does not rely on cultivation and is able to provide all the microbial genome contents[3]. However, the current mNGS methods are still facing several limitations, which hinder their large-scale applications[4, 5]. For example, the complexity of clinic specimens presents a big challenge for mNGS methods due to variable pathogen load, the presence of commensal microbial flora, and the high ratio of host : pathogen nucleic acids (up to 10^5^:1 in human sputum)[6]. Also, the current mNGS methods are mainly based on short-read sequencing platforms such as Illumina’s HiSeq technology, which require a library amplification procedure and is hardly to match the antimicrobial resistance (AMR) genes to individual bacterial species due to the short-read length[7].

Long read sequencing technologies, such as the Oxford nanopore technologies (ONT) and SMRT sequencing platforms, allow for fast, real-time sequencing of complex clinic samples[8, 9]. However, the lack of a library amplification procedure of the long-read sequencing technologies also leads to lower throughput compared to the short-read sequencing platforms, and is hard to detect pathogens at low copy. To overcome this drawback, several strategies including differential sample lysis methods, human DNA removal and microbial DNA enrichment methods have been developed and shown to improve the enrichment of microbial genome reads from complex samples[10-12]. Nevertheless, these enrichment approaches usually contain multiple steps, which might be compromised to complex clinic samples and are thus difficult for large scale clinic implementations[13].

Recently, the ONT platform introduced the nanopore adaptive sampling (NAS) method that enables real-time enrichment or depletion of molecules of interest from a mixed sample[14]. This approach enables target enrichment or depletion of unwanted DNA molecules (e.g., human DNA) directly during sequencing. While a DNA molecule is sequenced in the nanopore, the raw data is already compared in real time with references to decide whether the DNA molecule should be sequenced further (accepted) or removed directly from the pore (rejected)[15]. Each pore is individually manurable and can reverse the voltage on its pore to reject DNA molecules and sequence another one instead, increasing the sequencing capacity for molecules of interest[16]. The main advantage is that this depletion or enrichment method can be combined in addition to wet-lab depletion or enrichment methods and does not prolong the overall sequencing run time[17]. In the present study, we present a mNGS pipeline that features efficient selective ‘human host depletion’ NAS sequencing, and species identification and resistance genes prediction - ARGpore2. This pipeline can increase microbial sequencing yield from clinical respiratory samples and enables pathogen identification within 4.5 h.

## Materials and methods

### Ethics statement

The research was approved by the Ethical Review Committee of Huazhong University of Science and Technology Union Shenzhen Hospital (Nanshan Hospital) [Approval Number: KY-2020-032-01], and filed with the Ethical Committee of Southern University of Science and Technology [Approval Number: 20210131].

### Bacteria/Archaea culture

7 bacterial pathogens (*Acinetobacter baumannii, Escherichia coli, Enterococcus faecalis, Klebsiella pneumoniae, Pseudomonas aeruginosa, Serratia marcescens* and *Staphylococcus aureus*) and 2 archaea (*Haloferax mediterranei* and *Sulfolobus acidocaldarius*) were used in this study. Bacteria were grown overnight in 3 ml of Luria-Bertani (LB) broth in a 12 ml tube at 37 °C shaking at 200 rpm. Genomic DNA of each strain was extracted using QIAamp^®^ DNA Mini Kit (QIAGEN, Hilden, GERMANY) following manufacturer’s protocol. Gram-positive bacteria were pre-incubated with 20 mg/ml lysozyme for at least 30 min at 37°C. *Haloferax mediterranei* was cultivated at 37°C in nutrient-rich AS-168L medium (per liter, 150g NaCl, 20g MgSO_4_·7H_2_O, 2g KCl, 3g trisodium citrate, 1g sodium glutamate, 50mg FeSO_4_·7H_2_O, 0.36mg MnCl_2_·4H_2_O, 5g Bacto Casamino Acids, 5g yeast extract, pH 7.2). *Sulfolobus acidocaldarius* was cultivated in 6 mL of XTU medium (a xylose and tryptone [XT] medium supplemented with 0.02 g/L uracil (pH 3) at 75°C and 160 rpm with shaking. Genomic DNA of archaea was extracted using FastDNA^®^ Spin Kit for Soil (MP Biomedicals, USA) following manufacturer’s protocol.

### Human genomic DNA extraction

One blood sample of healthy people after health check in the Huazhong University of Science and Technology Union Shenzhen Hospital was collected. Human genomic DNA was extracted using QIAamp^®^ DNA Mini Kit (QIAGEN, Hilden, GERMANY) following manufacturer’s protocol for blood sample.

### Clinical sample collection

This study used respiratory samples from patients with suspected lower respiratory infections such as persistent (productive) cough, bronchiectasis, cystic fibrosis and exacerbation of chronic obstructive pulmonary disease (COPD, emphysema/chronic bronchitis)[18] in the Huazhong University of Science and Technology Union Shenzhen Hospital (Nanshan Hospital). The 11 Broncho-alveolar Lavage Fluid (BALF) samples were collected after cultivation and susceptibility testing at Microbiology Department and frozen at - 80°C until analysis. No patient identifiable information was collected. The only data collected were routine microbiology results, which detailed the pathogen(s) identified and their antibiotic susceptibility profiles (Supplementary Table 1).

### DNA extraction, quantification and quality control

DNA was extracted from 1ml of BALF samples using the ZymoBIOMICS (Zymo Research) Quick-DNA HMW MagBead kit per manufacturer’s instructions. DNA quantification was performed using the high-sensitivity dsDNA assay kit (Thermo Fisher, Q32851) on Qubit 4.0 Fluorometer (Thermo Fisher Scientific, US). DNA quality and fragment size were assessed using TapeStation 4150 (Agilent Technologies Inc, USA) automated electrophoresis platform with Genomic ScreenTape (Agilent Technologies, 5067-5365) and a DNA ladder (200 to >60,000 bp, Agilent Technologies, 5067-5366).

### Construction of a metagenomics mock sample and Nanopore sequencing

One metagenomics mock sample consisting of 7 bacterial and human genomic DNA were constructed based on Qubit measurements. The mock sample has 20% ‘target’ bacterial sequences with equal proportions of *A. baumannii, E. coli, E. faecalis, K. pneumoniae, P. aeruginosa, S. marcescens*, and *S. aureus*, while the rest 80% sequences were human genome. Approximately 1.5 μg of DNA mock was used to prepare a sequencing library using the Ligation Sequencing Kit (SQK-LSK109) according to manufacturer’s instructions. For library cleanup, long fragment buffer (LFB) was used to enrich DNA fragments of long sizes. Approximately 50 fmol of the prepared library was loaded onto R9.4.1 flow cell. Sequencing was performed using ONT GridION sequencing platform. NAS sequencing was applied using MinKNOW software, which allowed us to deplete human sequences dynamically. NAS sequencing and standard sequencing of the library were performed simultaneously on the same flow cell by setting the NAS channel from 1 to 256 (leave 257-512 channels for standard sequencing). The human reference genome GCA_000001405.28_GRCh38.p13 was used as the reference sequence, and host sequence depletion was enabled by selecting “Deplete”. The sample was sequenced by a standard 48-hour run script.

### Nanopore sequencing of clinical samples

In total, 11 samples were prepared and run on three ONT R9.4.1 flow cells. The first flow cell tested B1 sample spiked with 2 archaea (*H. mediterranei* and *S. acidocaldarius*). B1 sample genome was spiked with genomic DNA from 2 archaea at 1‰ total DNA by mass, and the mass ratio of *H. mediterranei* and *S. acidocaldarius* was 4:1. Library preparation and sequencing were the same as described above, NAS sequencing and standard sequencing for the library were performed simultaneously on the same flow cell by setting the NAS channel from 1 to 256 (leave 257-512 channels for standard sequencing). The second flow cell tested 5 samples including B2-B6, and the third flow cell tested 5 samples including B7-B11. Sequencing of 5 samples on a single flow cell using the Ligation Sequencing Kit (SQK-LSK109) with the ONT Barcoding Expansion Kit (EXP-NBD114). All steps were performed according to the Native barcoding genomic DNA ONT protocol, and set all the channels for NAS. Similarly, the human reference genome GCA_000001405.28_GRCh38.p13 was used as reference sequence, and host sequence depletion was enabled by selecting “Deplete”. These samples were sequenced by a standard 48-hour run script.

### Illumina sequencing of clinical samples

llumina sequencing libraries were prepared with 500 ng gDNA template for each sample according to the TruSeq DNA Sample Preparation Guide (Illumina, 15026486 Rev.C). Concentrations of the constructed libraries were measured using Qubit 4.0 and diluted to 1ng/μl. Agilent 2100 Bioanalyzer and Bio-RAD CFX 96 Real-Time PCR System (use Bio-RAD KIT iQ SYBR GRN) were used to qualify and quantify the sample libraries (library effective concentration > 10nM). The qualified libraries were then sequenced on Illumina Hiseq 2500 platform with 150 bp paired-end reads (Anoroad, Beijing, China).

### Human read removal

For Illumina sequencing data, the raw reads generated from samples were trimmed and filtered to remove low quality (Q□≤□20) and short reads (length < 50 bp) using Trimmomatic (version 0.39)[19]. Reads aligned to the human hg38 genome (GCA_000001405.28_GRCh38.p13) were removed (identity cutoff ≥□90%; maximum mismatches, 10□bp) by Bowtie2 (version 2.3.5.1)[20].

For Nanopore sequencing data, reads were basecalled using Guppy v4.2.2 GPU basecaller (Oxford Nanopore Technologies) during sequencing using the high accuracy basecalling model. Human reads were removed from basecalled FASTQ files using minimap2[21] to align to the human hg38 genome. Only unassigned reads were exported to a bam file using Samtools (-f 4 parameter)[22]. Non-human reads were converted back to FASTQ format using bam2fastx. These FASTQ files were processed for pathogen identification and antibiotic resistance gene detection with ARGpore2[23]. Further downstream analysis for genome coverage was performed using minimap2 with default parameters for long-read data (-a -x map-ont).

### Bacterial genome assembly

Genome assembly was performed first to remove reads shorter than 1,000 bp and with a mean quality score lower than seven. Filtered reads were aligned to a reference genome using minimap2 with default parameters for ONT long-read data (v.2.6-2.10)[21]. Finally, Canu was used to assemble mapped reads into contigs using this long-read sequence correction and assembly tool (v.1.6)[24, 25]. BLAST Ring Image Generator (BRIG) was used for BLAST comparisons of the genome assemblies generated[26].

### Pathogen identification and antibiotic resistance gene detection

The ARGpore2 antibiotic resistance genes (ARGs) pipeline was used for initial analysis of MinION data for the identification of bacteria present in the sample and any associated antimicrobial resistance genes. ARGpore2 supports the identification of bacteria, viruses, fungi, archaea and human reads. ARGpore2 utilizes Centrifuge, a k-mer-based read identification tool based on a Burrows-Wheeler transform and the Ferragina-Manzini index, to identify reads using the RefSeq database[27]. Relative abundance of a population was calculated as the number of reads assigned to this taxon in percentage of the total number of annotated reads could be assigned at a given taxonomic level.

ARGpore2 utilizes the nucleotide sequences of SARG database[28] for antibiotic resistance gene detection and identification by aligning input reads using LAST tool with custom settings as -a 1 -b 1 -q 2[29]. Next, tabular result of LAST search was filtered based on alignment length and similarity. Only alignments showing alignment length >50% of ARG length with similarity >60% were used for ARG counting[30]. If ARG-containing regions on Nanopore reads overlapped for more than 80% of the alignment length, only the best ARG hit (with the highest bit score) was kept for this overlapped region. This parameter currently reports resistance genes, acquired and chromosomal, but not resistance mutations/SNPs. Full manuals are publicly available for ARGpore2 on the GitHub (https://github.com/sustc-xylab/ARGpore2).

## Results

### Microbial enrichments from the mock community

We constructed a mock community to validate the NAS performance on enriching low abundance species. In the mock run, a flow cell was configured into two parts, where the 1-256 channels did NAS sequencing, and the 257-512 channels did normal sequencing as a control. The 48h sequence on the flow cell delivered 5.06 Gbp of effective long reads with an average read length of 2.18 kbp, while the NAS sequencing produced 1.99 Gbp reads with an average read length of 1.18 kbp, and normal sequencing produced 3.07 Gbp reads with an average read length of 4.86 kbp (Supplementary Table 2). In the NAS sequencing, the accepted sequence produced 1.14 Gbp reads with an average read length of 4.62 kbp, while the rejected produced 0.85 Gbp reads with an average read length of 592 bases.

More than 99.0% of the rejected reads and 24.5% of accepted reads were human genome sequences in the NAS sequencing. Whereas the normal sequencing, 89.6% reads were human genome sequences. This was a fairly ideal enrichment result, where microbial DNA got enriched from the mock sample is 882.63 Mbp in NAS sequencing, which is 2.7-fold compared with the microbial sequence from the normal sequencing approach (327.58 Mbp).

The microbial reads were next classified via the ARGpore2 pipeline to investigate their microbial and ARGs composition. The NAS method clearly showed very similar proportions to the control for 7 microorganisms, but all showed significant differences with the real proportions in mock sample (Fig. 1a). The reads were mapped against the microbial reference genomes via minimap2, counted via samtools depth v1.11 (bases sequenced per organism). For the mock sample, the NAS sequencing results provide sufficient depth (i.e., ≫30× coverage) to potentially assemble all 7 of the bacteria (Fig. 1b). The coverage of 7 bacterial genomes was lower (18× ∼ 32×) by normal sequencing results, potentially sufficient for assembly scaffolding (Fig. 1c). Significantly, 4 bacterial genomes reached 30 coverage depth in about four hours in NAS sequencing method (Fig. 1d).

**Fig. 1:**
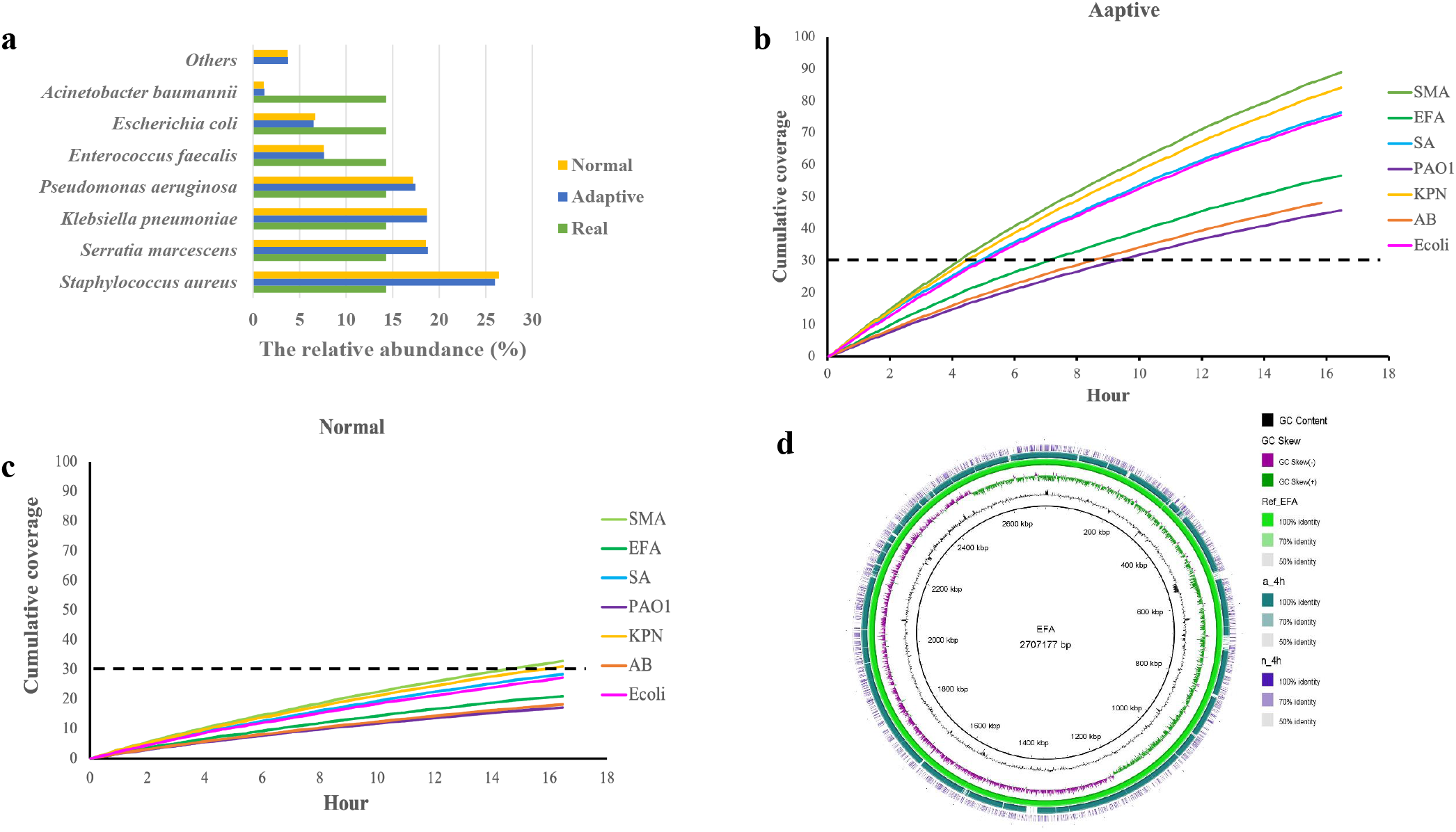
Enriching microbial species in mock community with NAS. a) Bar plot of the abundance of the seven microbial species in NAS and control runs. The NAS method clearly showed very similar proportions to the control for 7 microorganisms, but all showed significant differences with the real proportions in mock sample. (AB: *A. baumannii, E. coli*, EFA: *E. faecalis*, KPN: *K. pneumoniae*, PAO1: *P. aeruginosa*, SMA: *S. marcescens*, and SA: *S. aureus*) b) and c) The seven microbial species genome coverage of NAS (b) and normal sequencing (c). The NAS sequencing results provide sufficient depth (i.e., ≫30× coverage) to potentially assemble all 7 of the bacteria, but was lower (18× ∼ 32×) by normal sequencing results. d) *E. faecalis* genome assembly after 4 h of sequencing.

124 ARGs were identified from 4922 reads using the NAS sequencing, while 117 ARGs were identified from 1810 reads from the normal sequencing, and 109 ARGs were commonly detected by both methods (Fig. 2a). In NAS sequencing, 119 ARGs were inherent to 7 bacterial strains, whereas 112 ARGs could be assigned to a host strain in normal sequencing (Fig. 2b). Taking the *tet* gene family mediated tetracycline resistance as an example, 103 and 27 reads were aligned to *tet* genes using NAS sequencing and normal sequencing, respectively. *Tet* gene alignment was not seen within the first 5□min of normal sequencing, whereas two *tet* genes alignments were detected in the NAS sequencing by the same time point (Fig. 2c).

**Fig. 2:**
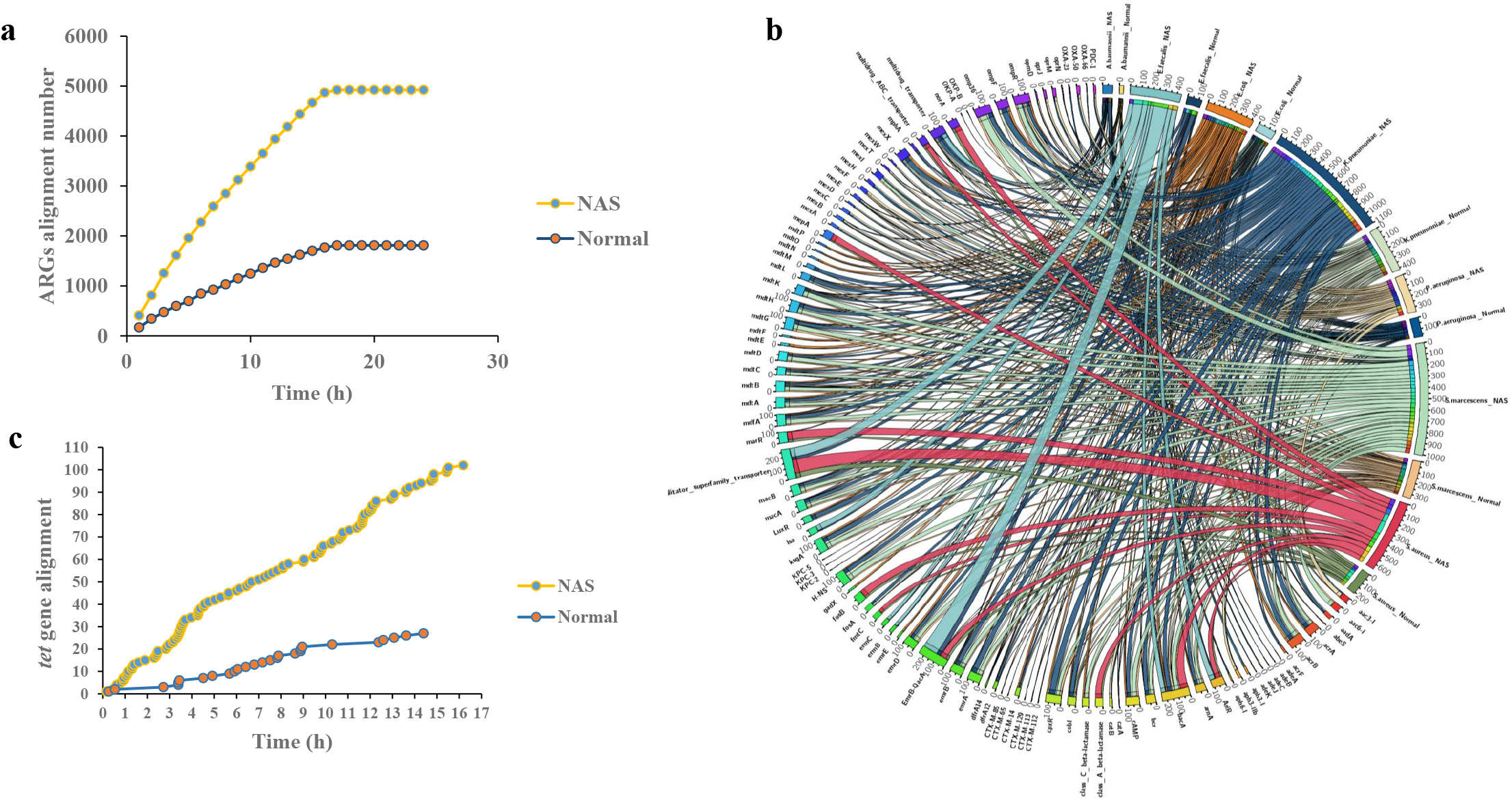
ARGs detection with NAS versue normal sequencing. a) ARGs alignment number of NAS versus normal sequencing. 124 antibiotic resistance genes were identified from 4922 reads using the NAS sequencing, while 117 ARGs were identified from 1810 reads from the normal sequencing. b) Chord diagram illustrating the correlations between ARGs and the 7 ARG-carrying pathogenic species. Thickness of the lines represents the number of samples observing such correlation. In NAS sequencing, 119 ARGs were inhere to 7 bacterial strains, whereas 112 ARGs could be assigned to a host strain in normal sequencing. c) *tet* genes alignment number of NAS versus normal sequencing. 103 and 27 tet genes were detected in NAS sequencing and normal sequencing, respectively. The tet gene was not detectable within the first 5□min of normal sequencing, whereas two tet genes alignments were detected in the aNASsequencing by the same time point.

### NAS sequencing application for one BALF sample

To evaluate whether the microbial enrichment using NAS sequencing is suitable for human BALF samples, we compared this method with a control experiment with normal ONT sequencing, as well as the Illumina sequencing using B1 sample.

The Illumina method resulted in 45.96 Gbp of sequencing data, while ONT normal and NAS sequencing yielded 6.49 and 1.30 Gbp of sequencing data, respectively. The yield of NAS sequencing was approximately 20.0% of normal ONT sequencing. After host contamination was removed from each read set, we observed an 8-fold enrichment of microbiome sequences using NAS on the ONT platform that normal and NAS sequencing resulted in 25.64 Mbp (0.39%) and 219.56 Mbp (16.44%) of microbiome sequence data, respectively (Supplementary Table 2). At the same time, Illumina sequencing resulted in 3.2 Gbp (2.76%) of microbiome sequence data.

To further examine the impact of sequencing methods on the metagenomic profiling, we included two species of marine Archaea (*S. acidocaldarius* : *H. mediterranei* = 4:1) to the B1 sample as internal reference. We compared the proportions of bacterial species of the experiments identified from the reads of the Illumina and Nanopore nomal/adaptive category to validate whether the overall microbial composition was retained. The *S. acidocaldarius* -to- *H. mediterranei* ratio was 3.6 :1 (reads 498 : 137) in the NAS sequencing, 2.6 :1 (reads 132 : 51) in the ONT normal sequencing, and *H. mediterranei* was undetected in the Illumina shotgun sequencing. Thus, the NAS sequencing had facilitated the accurate reporting of community composition, which could be partially retained by normal ONT sequencing, while poorly covered by Illumina short reads.

Sample sequenced by the Illumina method detected highest number of species (4791), followed by the ONT NAS (1559) and ONT normal sequencing (477) methods. Interestingly, the taxonomical profile of 10 species with the highest abundance showed similar profiles in all three methods. Although the pathogenic organisms reported by routine microbiology (*K. pneumoniae* and *A. baumannii*) were detected together in three methods. *A. baumannii* has a place in the top ten detected species of the three methods, but *K. pneumoniae* was found to be top ten only in the two ONT methods (Fig 3a). Worth noting is that species identification called in the first 1 h of the NAS sequencing run, provided 190-fold and 16-fold enrichment for *K. pneumoniae* and *A. baumannii* reads than the normal ONT sequencing (Fig 3b). The NAS sequencing maintained microorganism composition as the normal ONT experiments and significantly reduced the number of human reads, making it a robust choice for clinical mNGS approach for samples with high amounts of human genome DNA.

**Fig. 3:**
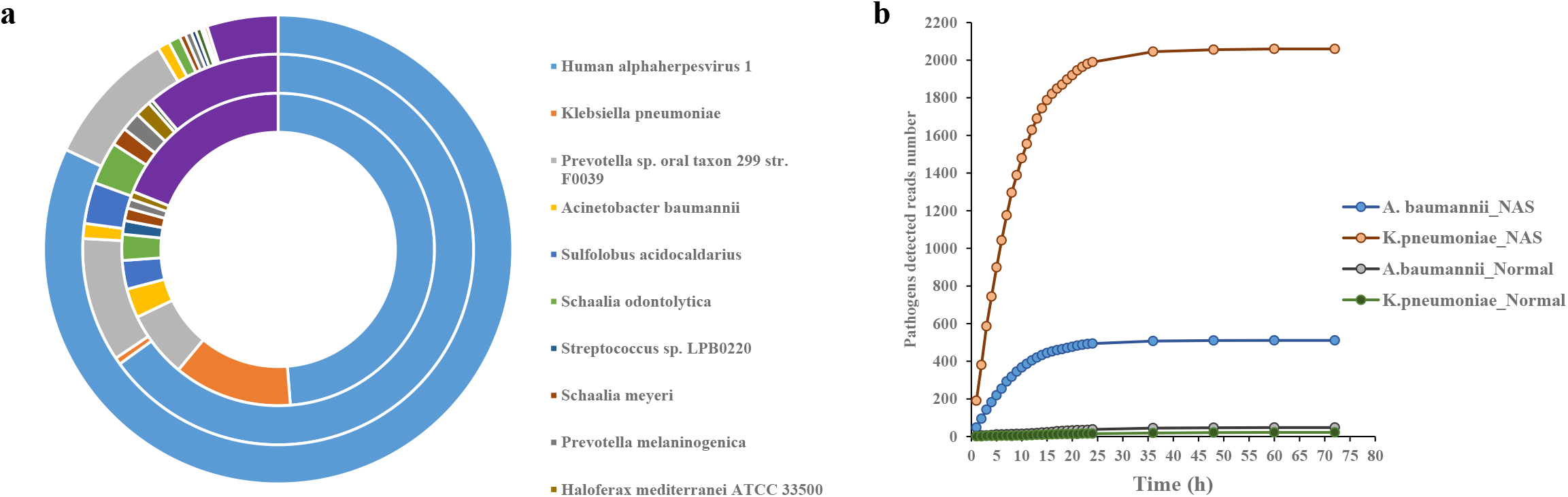
Species detection with NAS versue normal sequencing in B1 sample. a) Abundance of the top ten detected species with NAS, ONT normal and Illumina sequencing (the inner ring is NAS, central is ONT normal sequencing, the outer ring is Illumina sequencing). The taxonomical profile of 10 species with the highest abundance showed similar profiles in all three methods. *A. baumannii* has a place in the top ten detected species of the three methods, but *K. pneumoniae* was found to be top ten only in the two ONT methods. b) The pathogens(*K. pneumoniae* and *A. baumannii*) alignment number of NAS versus normal sequencing. In the first 1 h of the NAS sequencing run, provided 190-fold and 16-fold enrichment for *K. pneumoniae* and *A. baumannii* reads than the normal ONT sequencing.

In the B1 sample, 97 different ARGs (belonging to 15 different AMR types) were identified from 1503 reads of the NAS sequencing, while only 3 ARGs (belonging to 3 different AMR types) were identified from 211 reads of the normal ONT sequencing. Using the NAS sequencing method, *K. pneumoniae* was found to harbor 63 ARGs belonging to 15 AMR types, *A. baumannii* was found to harbor 31 ARGs belonging to 7 AMR types (Fig 4a). Using the normal ONT sequencing method, the ARGs carried by pathogens were not detected. In the first 1h, 2h, 3h of the NAS sequence run, 42 ARGs (belonging to 11 different AMR types), 67 ARGs (belonging to 13 different AMR types), 76 ARGs (belonging to 15 different AMR types) were detected, respectively (Fig 4b). In the 6h of the NAS sequence run, 7 AMR types (28 ARGs) and 15 AMR types (56 ARGs) were already found to be harbored by *A. baumannii* and *K. pneumoniae*, respectively (Fig 4c).

**Fig. 4:**
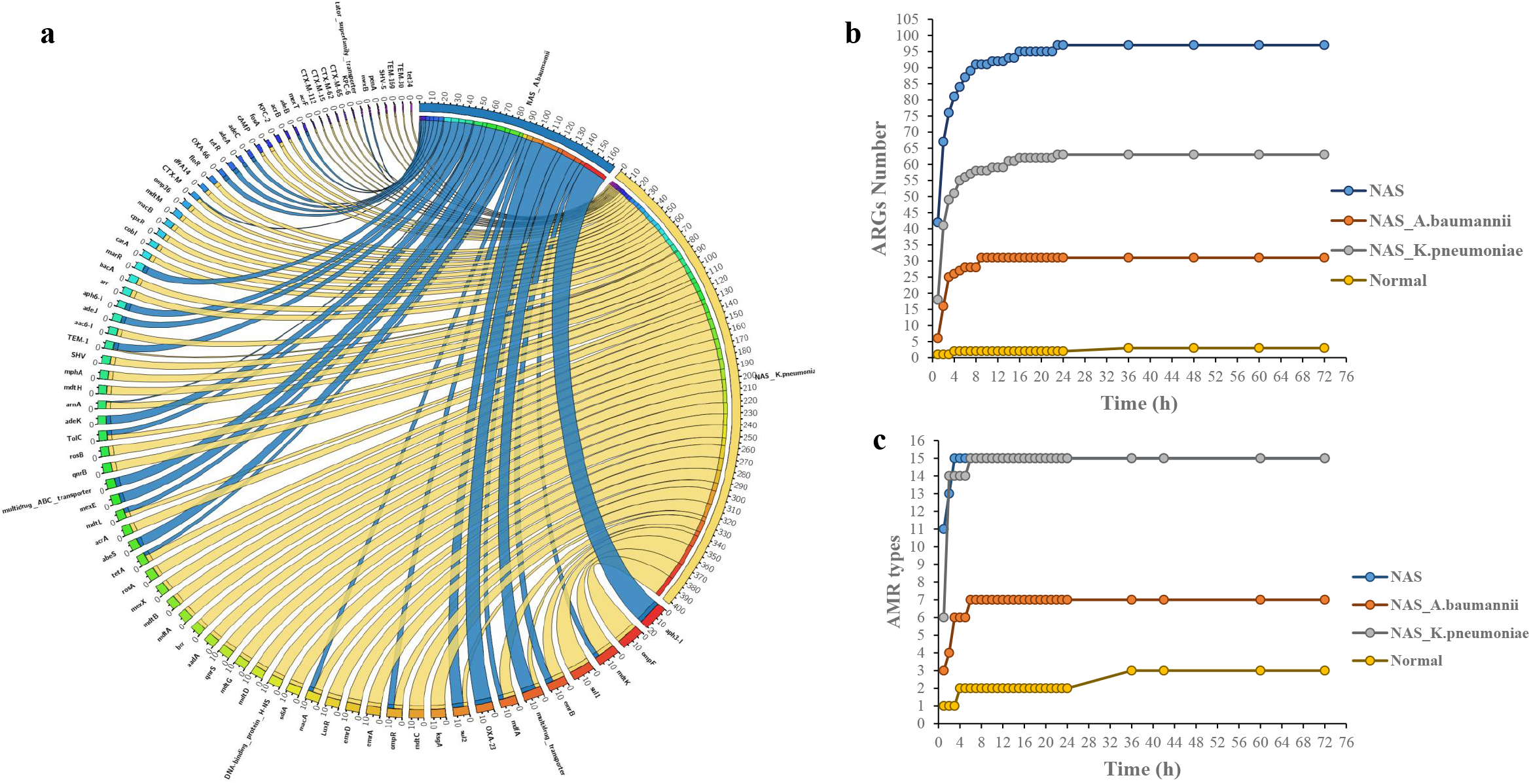
ARGs detection with NAS versue normal sequencing in B1 samples. a) Chord diagram illustrating the correlations between ARGs and the 2 ARG-carrying pathogenic species. Thickness of the lines represents the number of samples observing such correlation. In the NAS sequencing method, *K. pneumoniae* was found to harbor 63 ARGs belonging to 15 AMR types, *A. baumannii* was found to harbor 31 ARGs belonging to 7 AMR types. In the normal ONT sequencing method, the ARGs carried by pathogens were not detected. b) ARGs alignment number of NAS versus normal sequencing. c) AMR types detected number of NAS versus normal sequencing. In the 6h of the NAS sequence run, 7 AMR types (28 ARGs) and 15 AMR types (56 ARGs) were already found to be harbored by *A. baumannii* and *K. pneumoniae*, respectively.

### NAS sequencing for five BALF samples using a single MinION flow cell

In the B1 sample study, the yield of NAS sequencing was approximately 80% lower than that of normal ONT sequencing. This reduction in throughput can be partly attributed to the increased idle time of each nanopore caused by large numbers of ejections. We thus hypothesized that we could analyze multiple samples in real time on a single flow cell, to save cost and shorten the time of multi-sample detection.

We collected 10 BALF samples (B2-B11), per 5 BALF samples were sequenced on one flow cell using NAS sequencing method, and every sample was also sequenced using the Illumina platform. On average, Illumina sequencing experiments showed that each sample contains 99.27% (± 0.40%) human reads and 0.73% (± 0.44%) microbial reads. The ‘rejected’ fraction of the NAS experiments contained 99.56% (± 0.03%) human reads and 0.4% (± 0.03%) microbial reads, indicating a highly selective depletion process. Reads of the category ‘accepted’ contained low amounts of human reads (43.94% ± 16.47%) and high amounts of microbial reads (56.06% ± 16.47%) (Supplementary Table 2). Despite the incomplete removal of human fraction during NAS sequencing, the ‘accepted’ fraction of reads generally yielded more microbial reads compared to the control normal ONT experiments, underlining the capability of NAS to increase the microbial genome sequencing depth.

The microbial reads of ‘accepted’ fractions were classified via ARGpore2 pipeline to investigate their taxonomic and ARGs composition, while the Illumina sequencing reads were classified via centrifuge. Shannon alpha diversity analyses demonstrated that the samples sequenced by Illumina platform consistently resulted in a higher diversity than samples sequenced by NAS sequencing (Fig 5a). The pathogenic organisms reported by routine microbiology were detected in 7 samples: *S. pneumoniae* in B2 and B7, *P. aeruginosa* in B3 and B5, *S. aureus* in B4, *E. coli* in B8 and B9. There were three other mixed infections reported by routine microbiology, B6, B10 and B11, and both organisms were detected in all three samples using the NAS sequencing method. Except that *S. pneumoniae* was less detected in two samples (491 reads in B6, 358 reads in B7), other pathogens were detected in high abundance across the sample using the NAS sequencing method (Fig 5b). In the first 1 h of the NAS sequence run in B6 and B7, the numbers of sequencing reads detected to produce reliable pathogen identification were 16 and 13, respectively. In contrast, the first 1 h of normal ONT sequencing was unable to detect these pathogens.

**Fig. 5:**
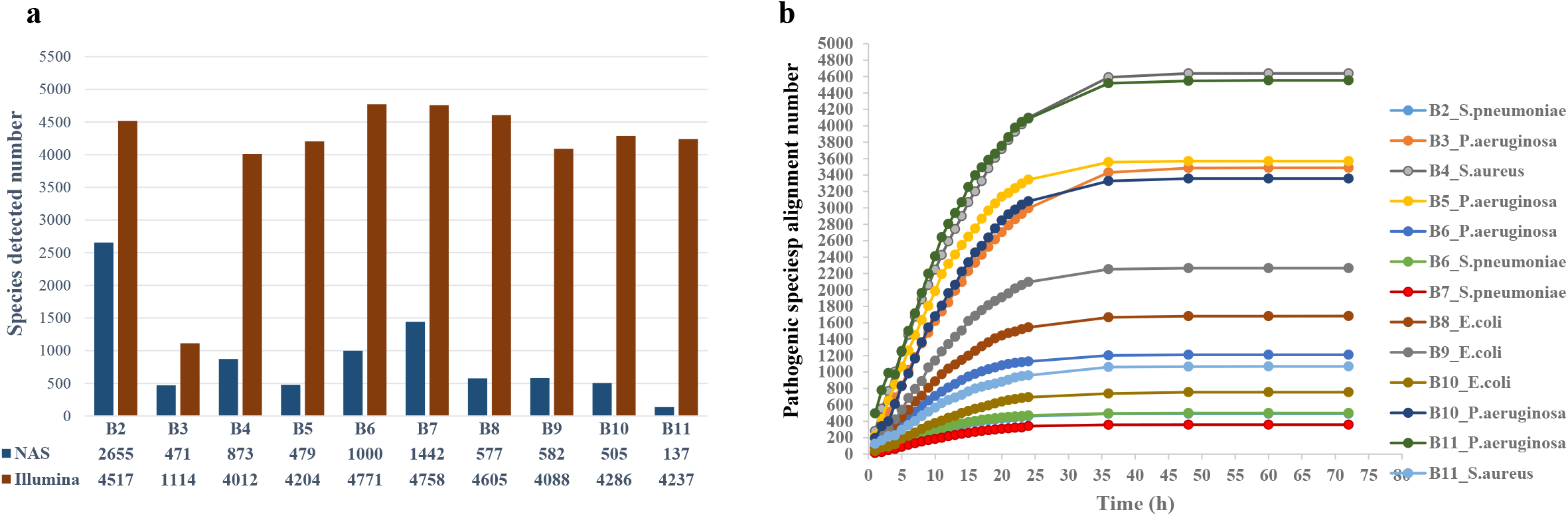
Species detection with NAS versue normal sequencing in B2-B11 samples. a) Species detected number with NAS and Illumina sequencing. Shannon alpha diversity analyses demonstrated that the samples sequenced by Illumina platform consistently resulted in a higher diversity than samples sequenced with NAS sequencing. b) The pathogens alignment number of NAS sequencing. Except that *S. pneumoniae* was less detected in the samples (491 reads in B6, 358 reads in B7), other pathogens were detected in high abundance across the sample using the NAS sequencing method.

NAS sequencing identified 135 ARGs belonging to 11 AMR types from the 10 BALF specimens. Among the 135 ARGs, 96 were harbored by the pathogenic species, for example, 11 ARGs were detected in *P. aeruginosa in B3 sample* (Fig 6a). A general consistency between detected genotype and phenotype experiment was observed (Supplementary Table 1) like *mecA* in MRSA (B4), *tetR* in tetracycline-resistant *E. coli* (B8, B9), *aac(3’)-II* in a tobramycin-resistant *E. coli* (B8), *mexA* in ceftazidime-resistant *P. aeruginosa* (B5, B11). There were 16 out of 135 ARGs whose associated resistance could not be confirmed due to limited phenotypic test in regular clinic practice; for example, *mac* genes were identified in several samples (B8, B9) but macrolide-lincosamide-streptogramin was not tested against the isolates cultured. A certain proportion of ARGs (39 out of 135) were likely originated from the normal flora since they were not detected from the pathogen genomes. Due to the inherent difficulty of isolating and sequencing all pathogens and commensal bacterial strains in a sample, the specificity and sensitivity of AMR detection by NAS sequencing could not be determined.

**Fig. 6:**
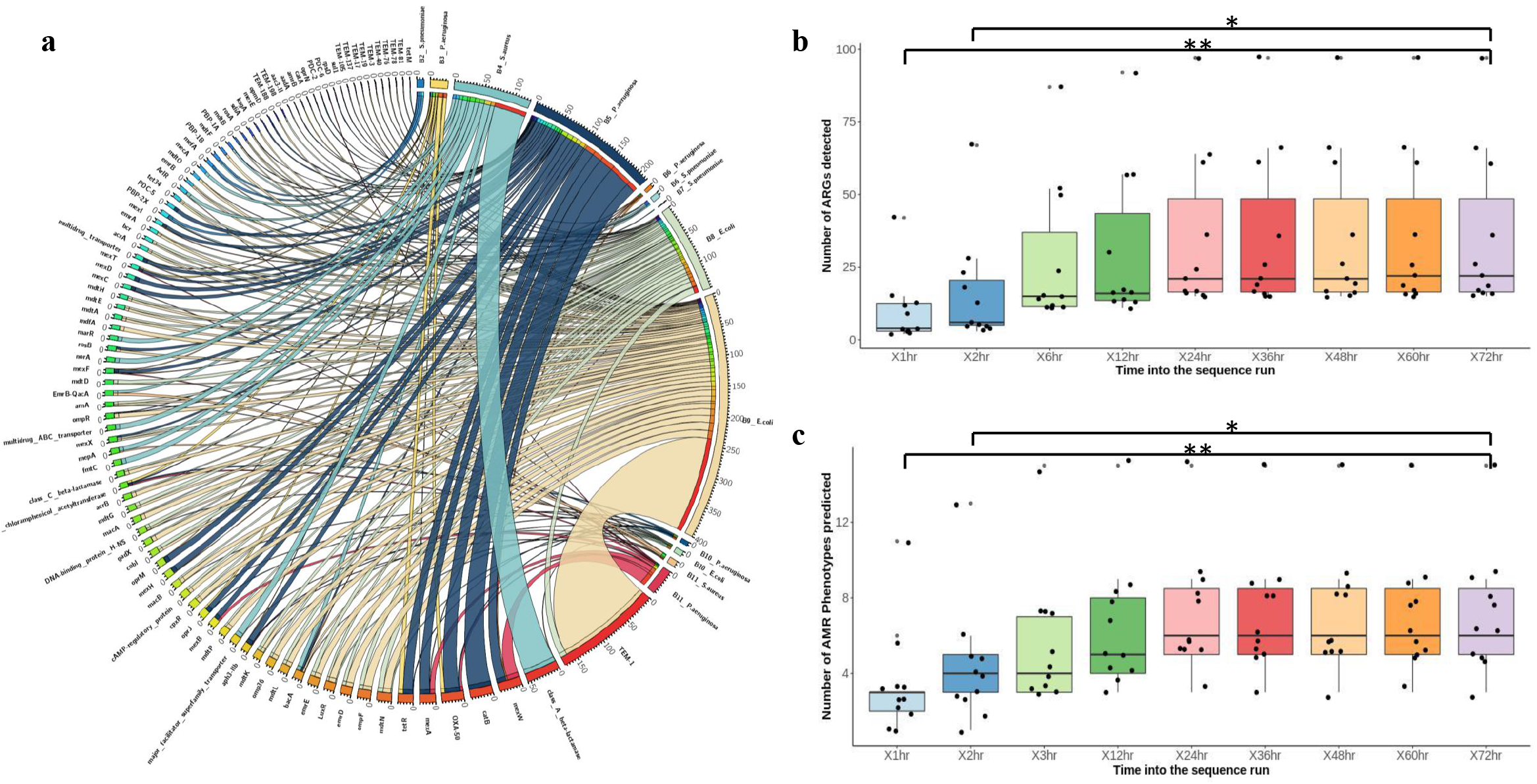
Detection of ARGs over the course of a 72-h sequence run. a) Chord diagram illustrating the correlations between ARGs and the ARG-carrying pathogenic species. Thickness of the lines represents the number of samples observing such correlation. NAS sequencing identified 135 ARGs belonging to 11 AMR types from the 10 BALF specimens. b) and c) To determine the optimum sequencing time required to detect ARGs and AMRphenotypes. The total number of ARGs (b) and AMR phenotypes (c) detected in each sample were plotted, and the Kruskal-Wallis H test was used to determine statistical significance in ARGs and AMR phenotypes detected at the different time points. Solid line, median; *, P value < 0.05; **, P value < 0.005.

### AMR prediction efficacy measurement

ONT sequencing allows for real-time analysis of sequencing data and the generation of multiple microbial diagnostic reports. A single report containing all diagnostic information, however, is preferable and reduces the likelihood of multiple empirical antimicrobials being administered. On average it takes 98 h for standard cultivation approach to complete species identification and AMR typing. In contrast, it takes a maximum of 3 h following sample collection to complete DNA extraction, Nanopore library preparation, and species identification and less than 1 h for AMR genotyping after the initiation of the NAS sequencing run, resulting in an average turnaround time of 4.5 h (Fig 7). Complete detection of microbial species and AMR genotyping varied depending on different types of samples used, while our experiment suggests that it takes 6 h of sequencing to complete the entire analysis (Fig 6b).

**Fig. 7:**
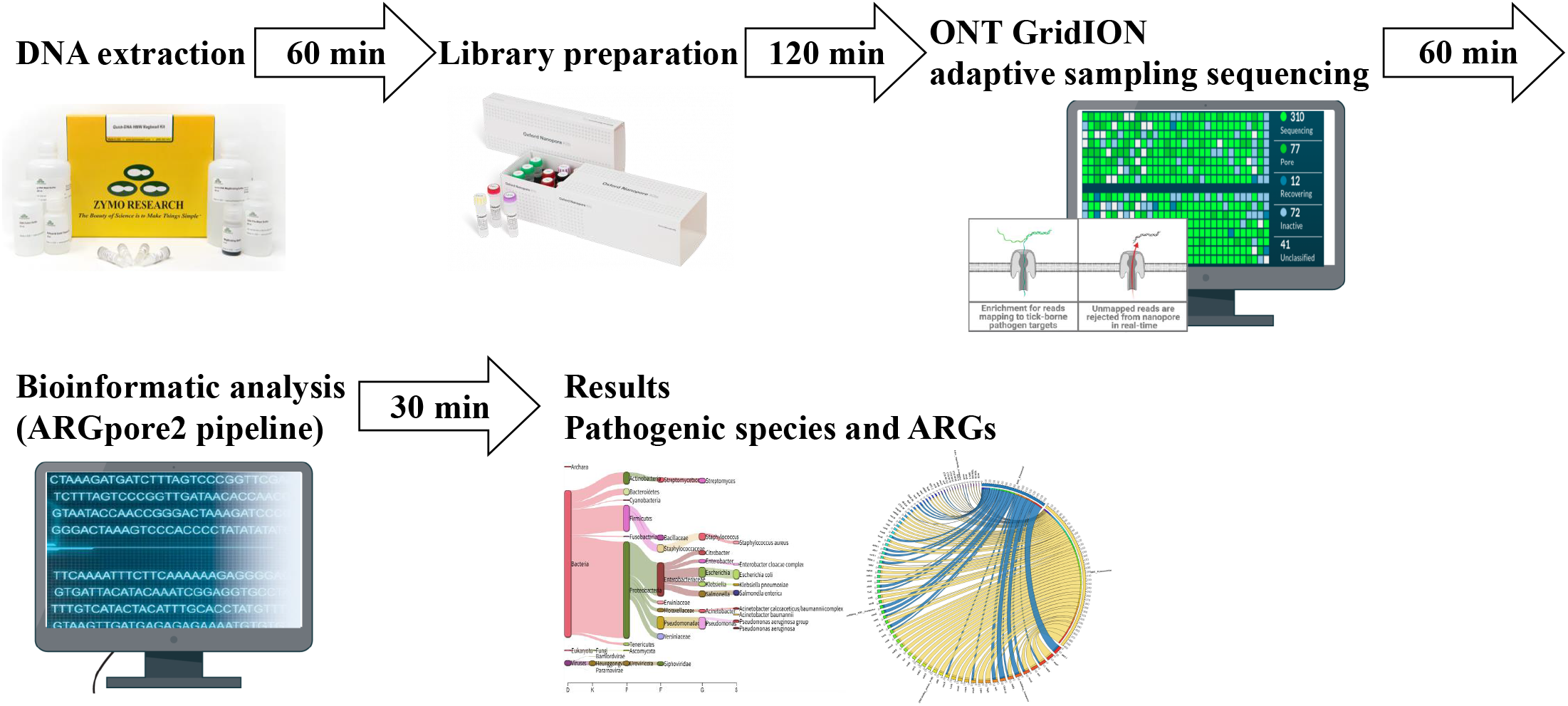
Schematic representation of the metagenomic pipeline. mNGS workflow includes microbial DNA extraction, library preparation, MinION NAS sequencing and ARGpore2 pipeline analysis. It takes a maximum of 3 h following sample collection to complete DNA extraction and Nanopore library preparation, and species identification with AMR genotyping could be performed within 1 h of initiating the NAS sequencing run, resulting in an average turnaround time of 4.5 h

The minimum sequencing run time required to produce reliable microbial and AMR identification was, therefore, investigated using sequenced data generated from the NAS method. Analysis of the number of bacterial reads detected revealed that it could reach the minimum detection threshold required to declare a sample to be microbial positive in the first 1 h of the NAS sequencing run. Identification of bacterial species was consistent at all time points, indicating that species identification could be called in the first 1 h of the sequencing run. In contrast, the number of detected ARGs and predicted AMR phenotypes increased over the course of the sequencing run. Time points 1 h and 2 h were associated with significantly reduced detection of ARGs and predicted AMR phenotypes. From 12 h onwards, there were no significant differences detected in both the number of ARGs and predicted AMR phenotypes. Comparison of AMR phenotypes prediction at different time points revealed that most AMR phenotypes were predicted in the first 6 h of the sequencing run (Fig 6b, 6c).

Overall, these results demonstrated that microbial identification can be determined within the first 1 h of the sequencing run, while the majority of the BALF ARGs can be characterized using a 6 h sequencing run.

## Discussion

The cultivation-based microbial diagnostics and susceptibility testing, in use for more than 70 years[31], have several limitations as guides for the appropriate clinical management of acute infections, mainly because of their slow sample-to-result turnaround[32, 33]. Rapid, accurate diagnostics would enable on-time treatment with appropriate antibiotics and improve treatment outcomes and antimicrobial stewardship alike[34]. We developed a novel mNGS dialogistic workflow, which combines the real-time sequencing data stream of NAS with automated analysis pipelines-ARGpore2, to shorten the turnaround time for simultaneously pathogen and ARG detection. Our mNGS workflow includes microbial DNA extraction, library preparation, GridION NAS sequencing and ARGpore2 pipeline analysis (Fig 7). This workflow increases the overall sequencing depth of bacterial sequences in clinic samples via ‘human depletion’ without changing the microbial composition during sequencing *per se*, which completes bacterial pathogen and ARGs detection in low respiratory specimens within 4.5 h.

Currently, several sequencing runs per sample or sequencers with higher throughput (e.g., PromethION in case of ONT) are necessary to increase the sequencing depth for metagenomic samples with low microbial material[35, 36]. Also, complicated wet laboratory methods have been developed to deplete the host DNA. These strategies all significantly increase the length and cost of diagnosis and thus hardly to be implemented clinically[10, 12, 37, 38]. Based on our mNGS workflow, DNA extracted from the BALF samples was sequenced in batches of 5 samples per flow cell. The total cost to perform DNA extraction, host DNA depletion, and sample sequencing using nanopore sequencing platform was approximately $267 per sample when sequencing was performed in a batch of 5 samples. The cost could be further reduced by bulk purchasing of the flow cells (R9), reducing price of flow cell from $1415 per cell to $1257 per cell, and the use of the rapid barcoding sequencing kit (SQK-RBK004), which enables multiple samples to be sequenced concurrently.

## Supporting information

Supplementary Table 1. The pathogenic organism and microbiology antibiogram results for BALF samples infected by Gram-negative and Gram-positive bacte

Supplementary Table 2. Detailed information of metagenomics data from 11 BALF samples

## Data Availability

All clinical sample sequence data (both Illumina and Nanopore) used in the present study has been deposited in the NCBI database under project ID PRJNA821338.

## Key Points

We developed a metagenomics workflow for ultra-sensitive diagnosis of bacterial pathogens and antibiotic resistance genes (ARGs) from clinic samples, 4.5h from sample to result.

The workflow includes microbial DNA extraction, library preparation, GridION NAS sequencing and ARGpore2 pipeline analysis.

This workflow increases the overall sequencing depth of bacterial sequences in clinic samples via ‘human depletion’ without changing the microbial composition.

This workflow displays 100% sensitivity and specificity for pathogen detection compared with cultivation method, and accurately detected antibiotic resistance genes at species level.

