## Supplementary Table 1. The pathogenic organism and microbiology antibiogram results for BALF samples infected by Gram-negative and Gram-positive bacte for "An ultra-sensitive bacterial pathogen and antimicrobial resistance diagnosis workflow using Oxford Nanopore adaptive sampling sequencing method"

**Supplementary Table 1.** The pathogenic organism and microbiology antibiogram results for BALF samples infected by Gram-negative and Gram-positive bacteria

| Sample | Organism cultured by microbiology | Gram Staining | Antibiogram |
|---|---|---|---|
| B1 | *Klebsiella pneumoniae* | Gram-negative | Amikacin S, Amoxicillin/clavulanic acid R, Aztreonam S, Ceftazidime S, Chloramphenicol S, Ciprofloxacin S, Gentamicin S, Levofloxacin S, Tetracycline R, Ceftriaxone S, Cefuroxime S, Colistin S, Ertapenem S, Imipenem S, Meropenem S, Minocycline S, Nitrofurantoin S, Norfloxacin S, Tegacyclin S, Tobramycin S |
| | *Acinetobacter baumannii* | Gram-negative | Amikacin R, Amoxicillin/clavulanic acid R, Aztreonam S, Ceftazidime S, Chloramphenicol S, Ciprofloxacin S, Gentamicin S, Levofloxacin S, Tetracycline R, Ceftriaxone S, Cefuroxime S, Colistin S, Ertapenem S, Imipenem S, Meropenem S, Minocycline S, Nitrofurantoin S, Norfloxacin S, Tegacyclin S, Tobramycin S |
| B2 | *Streptococcus pneumoniae* | Gram-positive | Oxacillin S, Penicillin S, Chloramphenicol S, Ciprofloxacin S, Fusidic acid S, Mupirocin S, Clindamycin S, Daptomycin S, Erythromycin S, Gentamicin S, Linezolid S, Minocycline S, Tetracycline S, Nitrofurantoin S, Rifampicin S, Tegacyclin S, Teicoplanin S, Vancomycin S |
| B3 | *Pseudomonas aeruginosa* | Gram-negative | Amikacin S, Gentamicin S, Ampicillin/sulbactam S, Cefoperazone/sulbactam S, Aztreonam S, Ceftazidime S, Cefatriaxone S, Cefepime S, Chloramphenicol S, Ciprofloxacin S, Levofloxacin S, Colistin S, Imipenem S |
| B4 | *Staphylococcus aureus* | Gram-positive | Oxacillin R, Penicillin R, Chloramphenicol S, Ciprofloxacin R, Fusidic acid R, Mupirocin S, Clindamycin S, Daptomycin S, Erythromycin S, Gentamicin R, Linezolid S, Minocycline S, Tetracycline S, Nitrofurantoin S, Rifampicin S, Tegacyclin S, Teicoplanin S, Vancomycin S |
| B5 | *Pseudomonas aeruginosa* | Gram-negative | Amikacin S, Gentamicin S, Ampicillin/sulbactam S, Cefoperazone/sulbactam S, Aztreonam S, Ceftazidime R, Cefatriaxone S, Cefepime R, Chloramphenicol R, Ciprofloxacin S, Levofloxacin S, Colistin S, Imipenem S |
| B6 | *Pseudomonas aeruginosa* | Gram-negative | Amikacin S, Gentamicin S, Ampicillin/sulbactam S, Cefoperazone/sulbactam S, Aztreonam S, Ceftazidime S, Cefatriaxone S, Cefepime S, Chloramphenicol S, Ciprofloxacin S, Levofloxacin S, Colistin S, Imipenem S |
| | *Streptococcus pneumoniae* | Gram-positive | Oxacillin S, Penicillin S, Chloramphenicol S, Ciprofloxacin S, Fusidic acid S, Mupirocin S, Clindamycin S, Daptomycin S, Erythromycin S, Gentamicin S, Linezolid S, Minocycline S, Tetracycline S, Nitrofurantoin S, Rifampicin S, Tegacyclin S, Teicoplanin S, Vancomycin S |

| | | | |
|---|---|---|---|
| B7 | *Streptococcus pneumoniae* | Gram-positive | Oxacillin S, Penicillin S, Chloramphenicol S, Ciprofloxacin S, Fusidic acid S, Mupirocin S, Clindamycin S, Daptomycin S, Erythromycin S, Gentamicin S, Linezolid S, Minocycline S, Tetracycline S, Nitrofurantoin S, Rifampicin S, Tegacyclin S, Teicoplanin S, Vancomycin S |
| B8 | *Escherichia coli* | Gram-negative | Amikacin S, Amoxicillin/clavulanic acid R, Aztreonam S, Ceftazidime I, Chloramphenicol S, Ciprofloxacin S, Gentamicin S, Levofloxacin S, Tetracycline R, Ceftriaxone S, Cefuroxime S, Colistin S, Ertapenem S, Imipenem S, Meropenem S, Minocycline S, Nitrofurantoin S, Norfloxacin S, Tegacyclin S, Tobramycin R |
| B9 | *Escherichia coli* | Gram-negative | Amikacin S, Amoxicillin/clavulanic acid R, Aztreonam S, Ceftazidime S, Chloramphenicol S, Ciprofloxacin S, Gentamicin S, Levofloxacin S, Tetracycline R, Ceftriaxone S, Cefuroxime S, Colistin S, Ertapenem S, Imipenem S, Meropenem S, Minocycline S, Nitrofurantoin S, Norfloxacin S, Tegacyclin S, Tobramycin S |
| B10 | *Pseudomonas aeruginosa* | Gram-negative | Amikacin S, Gentamicin S, Ampicillin/sulbactam S, Cefoperazone/sulbactam S, Aztreonam S, Ceftazidime S, Cefatriaxone S, Cefepime S, Chloramphenicol S, Ciprofloxacin S, Levofloxacin S, Colistin S, Imipenem S |
| | *Escherichia coli* | Gram-negative | Amikacin S, Amoxicillin/clavulanic acid S, Aztreonam S, Ceftazidime S, Chloramphenicol S, Ciprofloxacin S, Gentamicin S, Levofloxacin S, Tetracycline S, Ceftriaxone S, Cefuroxime S, Colistin S, Ertapenem S, Imipenem S, Meropenem S, Minocycline S, Nitrofurantoin S, Norfloxacin S, Tegacyclin S, Tobramycin S |
| B11 | *Pseudomonas aeruginosa* | Gram-negative | Amikacin S, Gentamicin S, Ampicillin/sulbactam S, Cefoperazone/sulbactam S, Aztreonam S, Ceftazidime R, Cefatriaxone S, Cefepime R, Chloramphenicol S, Ciprofloxacin S, Levofloxacin S, Colistin S, Imipenem S |
| | *Staphylococcus aureus* | Gram-positive | Oxacillin S, Penicillin S, Chloramphenicol S, Ciprofloxacin S, Fusidic acid S, Mupirocin S, Clindamycin R, Daptomycin S, Erythromycin R, Gentamicin S, Linezolid S, Minocycline S, Tetracycline S, Nitrofurantoin S, Rifampicin S, Tegacyclin S, Teicoplanin S, Vancomycin S |
